# Improving Labor Induction With Ultrasound: A Systematic Review

**DOI:** 10.1101/2025.01.30.25320795

**Authors:** Elisa Romero Callejas, Antonio Enamorado Plata, Miguel Ángel Luque Fernández, Juan Manuel Melchor Rodríguez, Miguel Ángel Montero Alonso

**Affiliations:** University of Granada; Department of Statistics and Operations Research, Faculty of Medicine. University of Granada; Department of Statistics and Operations Research, Faculty of Medicine. University of Granada, Instituto de Investigación Biosanitaria, ibs.GRANADA, Research “Unit Modelling Nature” (MNat), University of Granada

**Keywords:** Elastography, ultrasound, labor induction, systematic review

## Abstract

**Introduction:** Labor induction is performed when continuing the pregnancy poses a greater risk than ending it. Elastography is emerging as an increasingly relevant technique to predict the success of labor induction.

**Objective:** To evaluate the relevant literature regarding elastography as a predictive tool for the success of labor induction, resulting in an uncomplicated vaginal delivery.

**Methodology:** A systematic review was conducted on the predictive accuracy of elastography in labor induction, using PubMed, Scopus, and the Cochrane Library. A total of 10 studies were selected, covering the period from 2007 to 2022. The included studies were clinical trials and observational studies that analyzed elastography in predicting the success of induced labor. Success was defined as the completion of labor via vaginal delivery without maternal or neonatal complications.

**Results:** The elastography technique demonstrates an increased accuracy in predicting successful labor induction and vaginal delivery compared to the traditional use of Bishop score. Among other predictors of labor induction failure, defined as labor ending in a cesarean delivery, are: cervical length (Odds Ratio [OR] OR = 1.916; 95% Confidence Interval [1.451–2.530]), maternal age ≥35 years, and body mass index (BMI) ≥30 kg/m^2^ (OR = 2.257; 95% CI [1.353–3.767]). However, parity decreases the probability of labor induction failure (OR = 0.129; 95% CI [0.063–0.265]).

**Discussion:** The reviewed scientific evidence suggests that the use of elastography before induction improves the chances of a successful labor induction. Therefore, the right moment for the labor induction based on a clinical decision should be supplemented with other objective methods such as the elastography helping to evaluate the potential success of the induction and therefore, helping to improve maternal and infant health outcomes. Further research is recommended on the comparative effectiveness of different methods to identify the optimal time for labor induction, as well as to validate its clinical utility.

## INTRODUCTION

Labor induction involves stimulating uterine contractions before spontaneous labor begins, to achieve a vaginal delivery. It is usually recommended when continuing the pregnancy poses a greater risk than finishing it (add reference here). Even if the labor induction is recognized to be a safe technique, it could carry several risks for both the mother and the newborn baby. Therefore, proper indication in time, and risk assessment are crucial to reduce the likelihood of failure, possibly resulting in a cesarean delivery [1–3].

Labor induction has gained increasing relevance in recent years. The World Health Organization (WHO) recommends a population-based rate of 10% for induced labor, but many countries exceed this figure. In the United States, it surpasses 30% and is on the rise, while in Argentina and the United Kingdom, it reaches up to 20%, and in Canada, it stands at 21% (2,9,10).

Predictive factors for induction success or failure include gestational age, body mass index (BMI), parity, and the Bishop score. The Bishop score constitute the most used tool for the prediction of induction success or failure [10–12]. One of the main components of the Bishop score is the cervical ripening. Cervical ripening is key in predicting the success of labor induction, as an immature cervix can lead to a cesarean delivery in over half of the cases [3,13]. While the use of the Bishop score is simple, it is also subjective and prone to inter-observer variability, as it relies solely on the palpation of imprecise points. Therefore, it is necessary to establish easily applicable and reproducible quantitative objective diagnostic criteria to evaluate the ripening and consistency of the cervix. The use of ultrasound elastography has been used as complement to decide the right moment for the labor induction [13,14, 29].

We developed a systematic review of the literature aims to evaluate the scientific evidence to determine the comparative efficacy of different methods used to evaluate the success of failure of the labor induction, defined as the completion of labor via vaginal delivery without complications. We contrasted the use of the elastography, with the Bishop score and other predictive factors for the success or failure of labor induction.

## METHODS

This systematic literature review focuses on evaluating the predictive accuracy of elastography in determining the success of labor induction in pregnant women undergoing induction. Vaginal delivery is considered the success criterion for assessing the predictive capacity of elastography. We conducted an extensive search in the PubMed, Scopus, and Cochrane Library databases, covering the period from August 2007 to January 202. We used as keywords: Labor Induction, Elastography, Ultrasound, Vaginal Delivery, and Bishop Score, while combining them with links like AND as well as OR. Following the search criteria and the selection process, a total of 182 studies were identified. The inclusion criteria were controlled clinical trials, as well as prospective and retrospective observational studies, investigating the use of elastography as an evaluation method in pregnant women undergoing labor induction. Studies that did not include vaginal delivery as the success indicator for labor induction were excluded. No language restrictions were applied. Figure 1 shows the flow of articles included in this review.

**Figure 1:**
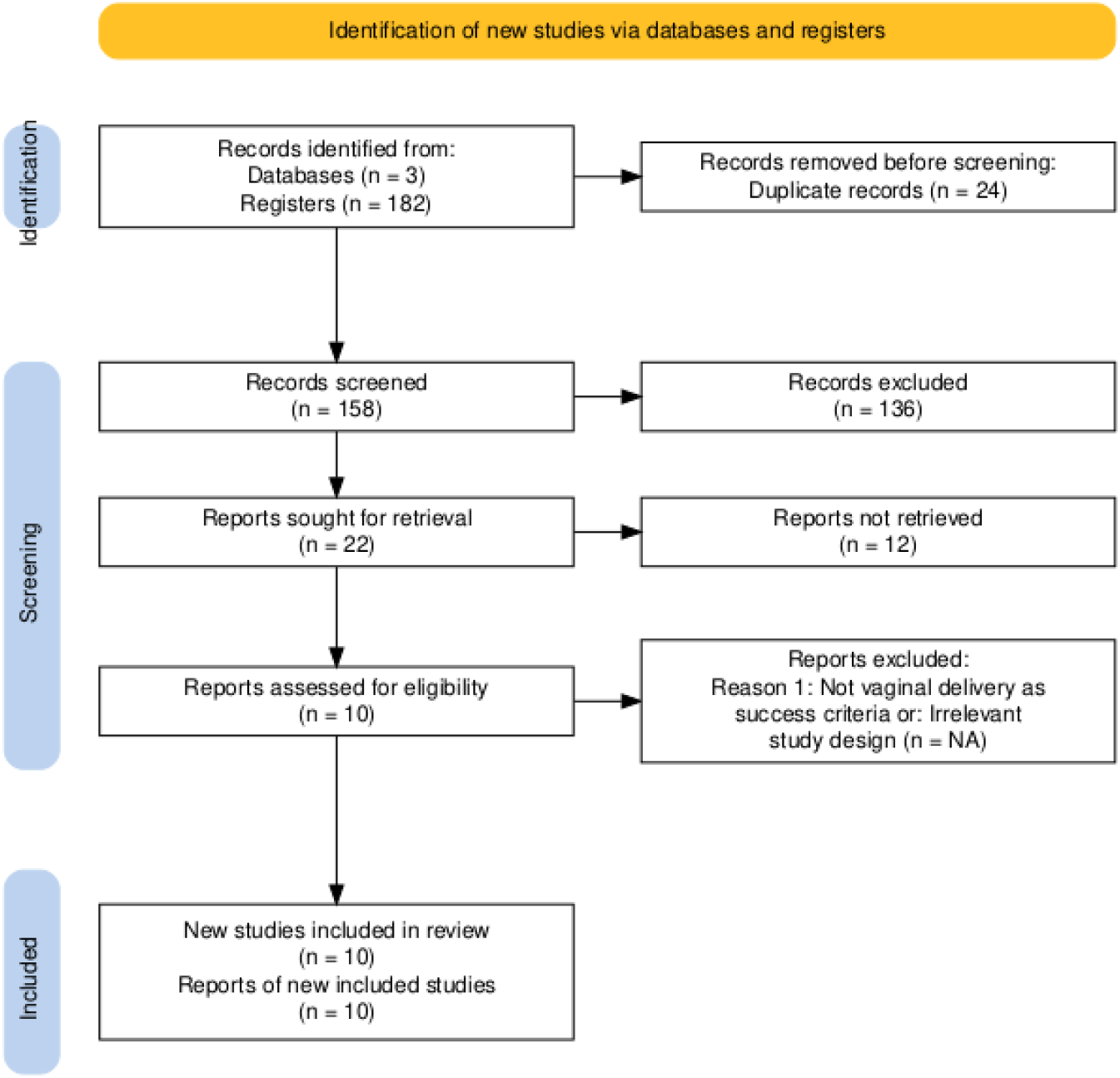
Flowchart of the search and selection of the included literature. PRISMA Flow Diagram.

Using the identified articles, we conducted a double-blind selection of articles to identify a total of 10 articles meeting the inclusion criteria for the review. 24 articles were excluded due to duplication. Of the remaining 158 articles, 136 were eliminated after reviewing the title, and 12 were excluded after reading the abstract [14,16–24]. From the 10 articles finally selected, relevant data were extracted, including the year of publication, the statistical methodology used, sample size, population characteristics, variables analyzed, study objectives, results obtained, conclusions, and methodology employed.

## RESULTS

Table 1 shows the count of the occurrence of the most relevant variables mentioned above in the 10 studies selected for this review. The variables “Parity” and “Cervical Length” are the most frequently key words mentioned in the selected studies.

**Table 1:** Number of studies by the most frequent key words

| Variable | Number of studies |
| --- | --- |
| Parity | 5 |
| Cervical length | 7 |
| Bishop score | 4 |
| Gestational age | 1 |
| Maternal age | 1 |
| BMI at the time fo delivery | 1 |

Table 2 presents all the extracted information from each of the 10 selected studies. In all these studies, the utility of cervical elastography for assessing and predicting the success of labor induction is emphasized, particularly in comparison to traditional methods such as the Bishop score and the cervical length measurement. Overall, these studies analyze the variables that can aid in predicting the failure or success of the induction technique.

**Table 2:** Summary of the extracted information from each selected article, n=10

| STUDY/YEAR<br>OF<br>PUBLICATION | METHODS | SAMPLE | VARIABLES | RESULTS |
| --- | --- | --- | --- | --- |
| Preliminary Results on the Preinduction Cervix Status by Shear Wave Elastography (20) **2022** | **PROSPECTIVE OBSERVATIONAL STUDY**<br>**Correlation analysis:** before and after maturation (SWS) / Bishop. Spearman rank correlation.<br>**Mann-Whitney U test;** for categorical variables, the Chi-square test was used.<br>**Wilcoxon signed-rank** test in R to compare before and after maturation.<br>**F test of likelihood ratio** with a significance level of 0.05.<br>95% CI. Two-tailed p-value of 0.05 for all tests. | **n=54** | Maternal age (years)<br>GA (gestational age) (weeks)<br>BMI (Body Mass Index) before pregnancy (kg/m<sup>2</sup>)<br>BMI at the time of delivery (kg/m<sup>2</sup>)<br>Newborn weight (g)<br>p=≤0.01<br>Nulliparous<br>Previous cesarean delivery<br>p=≤0.01<br>SWS (Shear Wave Speed) external operating system (EM)<br>p=≤0.01<br>SWS internal operating system (EM)<br>Cervical SWS box (EM)<br>p=≤0.05<br>CL (cervical length) (mm)<br>p=≤0.05<br>Bachelor's degree | **Prevalence of nulliparous women, with a lower Bishop score and a longer cervix in the cesarean group.**<br>**Objective:** To evaluate the preliminary potential capability of SWE (Shear Wave Elastography) to provide quantitative data and objective information about the mechanical state of the cervix through SWS (Shear Wave Speed) prior to labor induction.<br>**Conclusion:** The main findings revealed that the SWS of the external cervical os and cervical length were relevant in predicting induction failure. This technology is a good predictor of success in labor induction. |
| The predictive value of cervical shear wave elastography in the outcome of labor induction (18)<br>**2020** | **PROSPECTIVE STUDY**<br>**Multivariate regression analysis Kolmogorov-Smirnov test** to assess the normality of the variables.<br>Continuous variables with a normal distribution were compared using the **Student's t-test**.<br>Parameters with a non-normal distribution were compared using the **Mann-Whitney U test**.<br>Categorical variables were compared using the **Chi-square test** or **Fisher's exact test**.<br>Intra- and inter-observer reproducibility was evaluated using the **intraclass correlation coefficient (ICC)** and **Bland-Altman plots**.<br>SWE values between different cervical regions were compared using the **Wilcoxon signed-rank test**.<br>**Conditional stepwise elimination method** to generate the regression model and determine independent predictors.<br>The **area under the curve (AUC)** was compared using the **De Long test**. | **n=475** | <u>Odds ratio (CI 95%)</u><br>maternal age ($\geq 35$ years old) (p-value 0.023)<br>Materna height (cm)<br>(p=0.001)<br>OR=0.907 (0.864-0.952)<br>BMI at the moment of delivery ($\geq 30$ kg/m<sup>2</sup>)<br>(p=0.002)<br>OR=2.257 (1.353-3.767)<br>Multiparous<br>(p=0.001)<br>OR=0.129 (0.063-0.265)<br>EFW (estimated fetal weight) (g)<br>(p=0.023)<br>OR=1.001 (1.000-1.001)<br>Bishop score<br>(p=0.001)<br>OR=0.605 (0.515-0.71)<br>Cervical Length (cm)<br>(p=0.001)<br>OR=1.916 (1.451-2.530)<br>Posterior cervical angle (°) (p-value 0.04)<br>Progression angle (°)<br>(p=0.001)<br>OR=0.953 (0.931-0.974)<br>Internal cervical wave elastography (SWE)<br>(p=0.001)<br>OR=1.43 (1.214-1.684) | Vaginal delivery n=393 (82.7%)<br>Cesarean section n=82 (17.3%)<br>Indications<br>Not entering active phase (n=40)<br>Fetal distress (n=20)<br>Lack of progression (n=19)<br>Other (n=3)<br>**Objective:** To evaluate whether cervical SWE can improve the predictive performance of the outcome of labor induction when combined with other ultrasound-based assessments compared to the Bishop score alone.<br>**Conclusion:** Shear wave elastography is a useful tool in the pre-labor induction evaluation of cervical stiffness. The combination of ultrasound cervical length and shear wave elastography is superior to the Bishop score in predicting labor induction failure. |
| Quantitative Elastography of the Cervix for Predicting Labor Induction Success (17)<br>**2015** | **PROSPECTIVE STUDY**<br>Multivariate analysis (dependent variable: induction failure)<br>**Spearman correlation test** to evaluate the correlations between cervical ST and functional cervical length.<br>**Kruskal-Wallis and one-way ANOVA tests** for variance analysis. | **n=77** | **Failed labor induction vs successful labor induction.**<br>**Maternal age at the time of delivery** (years)<br>**BMI before pregnancy** (kg/m<sup>2</sup>)<br>**Gestational age at the time of delivery** (weeks). p=< 0.05<br>**Neonatal weight** (grams)<br>**Bishop score**<br>**Cervical length** (mm)<br>**Functional cervical length** (mm). p=<0.05<br>**TS (tissue stiffness)**. p=<0.05<br>**Use of prostaglandins** (mg). p=<0.05 | **Objective:** To evaluate the diagnostic accuracy of assessing cervical tissue stiffness (TS) to predict a successful labor induction.<br>**Conclusion:** Preliminary data show the potential utility of quantitative cervical elastography in predicting the failure of labor induction. |
|  | **Chi-square test** for the comparison of multiple categorical variables.<br>**Wilcoxon test, t-test** for continuous variables, **Chi-square test** and **Fisher's exact test** for categorical variables in bivariate analysis.<br>**ROC curves (Receiver Operating Characteristics)** to assess the predictive accuracy of outcomes in each diagnostic method studied.<br>AUCs were compared using the **De Long test** for two correlated ROC curves. |  | **Time from induction to delivery** (hours), $p < 0.05$<br>**Time from induction to delivery** (hours), $p < 0.05$ |  |
| Successful induction of labor: prediction by preinduction cervical length, angle of progression and cervical elastography (21) **2014** | **PROSPECTIVE STUDY**<br>**Regression analysis.**<br>**Logistic regression analysis:** which variables were significant predictors of vaginal delivery.<br>**Regression analysis:** to determine which of the aforementioned characteristics, besides the mode of delivery, were significant predictors of the square root-transformed induction-to-delivery interval.<br>**Student's t-test or Mann-Whitney U test** for continuous variables.<br>**Chi-square tests or Fisher's exact test** for categorical variables. | n=99 | **<u>Odds ratio (CI del 95%)</u>**<br>Age<br>Materna Weight (kg)<br>Materna Height (cm)<br>GA<br>RACE<br>**Nulliparous**<br>$P=0.001$<br>OR=0.106 (0.023–0.481)<br>**Bishop score**<br>$p=0.037$<br>OR=1.274 (1.020–1.592)<br>**Cervical length (mm)**<br>$p=0.049$<br>OR=0.955 (0.913–1.000)<br>Progression angle (PA)<br>Elastography score<br>Epidural analgesia<br>Occipital position<br>**Interval from induction to delivery (min)**<br>$p=0.001$<br>Neonatal Weight at birth (g)<br>Apgar score at 5 min < 7<br>**Admission to the neonatal unit**<br>$p=0.039$ | **Vaginal delivery n=66 (66.7%)**<br>**Cesarean section n=33 (33.3%)**<br>**Fetal distress n=7**<br>**Lack of progression n=26 (78.8%)**<br>**<u>Objective:</u>** To examine the potential value of cervical length prior to induction, cervical elastography, and the progression angle (PA) in predicting successful vaginal delivery and the interval between induction and delivery.<br>**<u>Conclusion:</u>** In women undergoing labor induction, the PA and elastography score at the internal os are unlikely to be useful for predicting vaginal delivery and the interval between induction and delivery.<br>There are no significant differences in the PA and elastography score at the internal os between women with vaginal delivery and cesarean section.<br>There are significant associations between cervical length and both the PA and elastography score. |
|  |  |  |  | 3. The PA and elastography score do not improve the prediction of vaginal delivery and the interval between induction and delivery provided by parity and cervical length. |
| Quantitative sonoelastography of the uterine cervix prior to induction of labor as a predictor of cervical dilation time (22)<br>**2014** | **CROSS-SECTIONAL STUDY**<br>LINEAR REGRESSION<br>Analysis of variance (ANOVA).<br>Receiver Operating Characteristic (ROC) analysis.<br>Intraclass correlations (ICC). | n=49 | **Maternal age (years) (mean) BMI (kg/m<sup>2</sup>) (mean) Parity (%): Multiparous women had a significantly shorter mean dilation time (min) than nulliparous women**<br>- Nulliparous: p = 0.04<br>- Multiparous: p =< 0.01<br>**Gestational age at the time of measurement (days) (mean)**<br>The **Young's modulus** is a parameter that characterizes the behavior of an elastic material according to the direction in which a force is applied. It is one of the most widely used methods for assessing the elasticity of a material. | The **approximate Young's modulus** was associated with cervical dilation time during active labor (p= < 0.01).<br>The figures for the **Bishop score** (p = 0.37).<br>**Quantitative cervical elastography** (sensitivity 74% and specificity 69%) has better results for predicting prolonged cervical dilation time than the Bishop score (sensitivity 53% and specificity 46%) and cervical length (sensitivity 66% and specificity 54%).<br>Additionally, we found an **intraclass correlation coefficient (ICC)** of 58% between observers using this method.<br>**Objective:** To evaluate the association between the Young's modulus assessed before labor induction and cervical dilation time, and to assess the approximate Young's modulus as a predictor of prolonged cervical dilation time.<br>**Conclusion:** The Young's modulus is superior to the Bishop score and cervical length in predicting dilation time and the risk of prolonged dilation time after induction. |
| Predictive value of cervical length by ultrasound and cervical strain elastography in labor induction at term (23)<br>**2021** | **PROSPECTIVE OBSERVATIONAL STUDY**<br>**Median (interquartile range) and number (%)**<br>**Intraclass correlation (ICC) and the 95% confidence interval (CI)**<br>**Mann-Whitney U test** for continuous variables and the **chi-square test** or **Fisher's exact test** | n=73 | **Maternal age**<br>**GA**<br>**Severity**<br>**Weight gained**<br>**Pre-pregnancy BMI**<br>- (P=0.04)<br>**BMI**<br>- (P=0.017)<br>**Indications for IOL:**<br>- Prolonged pregnancy<br>- Gestational diabetes<br>- Hypertensive disorder<br>- Cardiotocography | **Two groups**<br>**Vaginal delivery <24h**<br>**Vaginal delivery >24h**<br>The **BMI** was significantly higher in the group with the longer time interval (>24 hours) (P = 0.045).<br>In the ≤24-hour group, the median **CL** was significantly shorter (P = 0.005) and the **Bishop score** was higher (P = 0.025).<br>**Objective:** To examine whether the addition of |
|  | for categorical variables.<br>The **area under the curve (AUC)** of the prediction model was calculated using a **receiver operating characteristic (ROC)** curve. |  | – Oligohydramnios<br>– Thrombophilia<br>– Cervical length<br>**Cervical length**<br>• (p=0.044)<br>EIC (elasticity contrast index)<br>HR (hardness ratio)<br>**IOS (mean stiffness level of the internal operating system)**<br>- (P=0.033)<br>EOS (mean stiffness level of the external operating system)<br>Ratio<br>**Bishop score**<br>- (p=0.031)<br>**GA at the time of delivery**<br>**Neonatal weight at birth**<br>**Neonatal sex**<br>**Delivery hemorrhage** | cervical elastographic parameters measured by **ElastoScan** for the cervix (**E-cervix**) improves the predictive value of **cervical length (CL)** in term labor induction.<br>**Conclusion:** Measurement using **E-cervix** is relatively reproducible. The addition of cervical tension elastography slightly improves the predictive performance of CL in vaginal delivery within 24 hours.<br>1. The measurement of transvaginal CL and elastography using **E-cervix** were fairly reproducible.<br>2. There were significant differences in CL and the **Bishop score** between women who achieved uterine contractions within 24 hours and those who did not.<br>3. There were significant differences in CL, **HR**, and **IOS** between women who achieved vaginal delivery within 24 hours and those who did not.<br>4. The addition of HR and IOS slightly improved the predictive performance of vaginal delivery within 24 hours using CL. |
| Sonographic cervical assessment to predict the success of labor induction: a systematic review with metaanalysis (15)<br>**2007** | **EVALUATION OF PROSPECTIVE TRIALS**<br>**META-REGRESSION ANALYSIS** with cut-off values, year of study, and location as covariates.<br>Odds ratios (95% CI) were calculated for each outcome for the | 20 trials<br>n=3101 | **Odds ratio (IC 95%): comparison of CL and Bishop score:**<br>**Successful induction**<br>- **OR=0.82 (0.37-1.80)**<br>**Vaginal delivery**<br>- **OR=0.72 (0.17-2.98)**<br>**Vaginal delivery in less than 24h**<br>- **OR=0.59 (0.28-1.23)**<br>**Active labor**<br>- **OR=0.82 (0.27-2.54)** | **Objective:** To review the literature evaluating ultrasound cervical assessment to predict a successful labor induction.<br>**Conclusion:** Ultrasound cervical length was not an effective predictor of a successful labor induction. Additional assessment of cervical ripening in predicting labor induction appears to be warranted. |
|  | transvaginal ultrasound group compared to the Bishop score group for selected outcomes. |  | There are no significant differences between the diagnostic accuracy of cervical length or the Bishop score in predicting a successful labor induction. |  |
| Elastography of the uterine cervix: implications for success of induction of labor (19)<br>**2011** | The normality of the variables was tested using the **Kolmogorov-Smirnov test**, and the means were compared using the **Student's t-test**.<br>The **Pearson correlation coefficient** was used to assess the relationship between the IE of different parts of the cervix and the Bishop score as well as the measurement of cervical canal length before labor induction. | **n=29** | Maternal Age<br>GA at the time of the examination<br>**Parity**<br>**p=0.01**<br>**Bishop score**<br>**p=0.04**<br>Cervical Length in mm<br>*Elastography index*<br>**Inner operation system**<br>**p=0.024**<br>Cervical canal<br>Extreme OS | **18 labor inductions: FAILURE = 16 patients (55.2% of women) SUCCESS = 13 (44.8%) 9 women (69.2%) delivered vaginally 4 women (30.8%) underwent cesarean section.**<br>The indications for cesarean section were fetal asphyxia in three cases and threatened uterine rupture in one patient after a previous cesarean.<br>**Objective:** To conduct preliminary research on the use of elastography for cervical assessment to determine the effectiveness of this method for evaluating cervical consistency.<br>**Conclusion:** Cervical elastography may be an objective method for assessing tissue softening in the area of the internal os before labor induction. |
| Diagnostic accuracy of cervical elastography in predicting labor induction success: a systematic review and meta-analysis (24)<br>**2016** | **SYSTEMATIC SEARCH, REVIEW, AND META-ANALYSIS OF PROSPECTIVE OBSERVATIONAL STUDIES**<br>The **Cochran Q test** was used to evaluate heterogeneity among the studies.<br>The **inconsistency index** was used to detect the percentage of variability due to heterogeneity rather than sampling errors.<br>To assess the presence of publication bias, the **Egger test** was utilized (considering a P value < 0.05 as statistically significant) along with funnel plots for asymmetry. | 6 studies<br>n=323 | **successful induction results in the reviewed articles**<br>**Cervical Elastography**<br>OR (CI 95%)<br>2.479 (1.184–5.19)<br>– Hwang et al. 2013. Elastographic index (25)<br>6.279 (2.662–14.813)<br>– Hwang et al. 2013. Cervical hard area<br>1.944 (0.719–5.257)<br>– Fruscalzo et al. 2014. Cervix TS (26)<br>6.5 (1.602–26.375)<br>– Hee et al. 2014 Young's modulus (21)<br>group estimation: 3.501 (1.930–6.349)<br>**Cervical Length**<br>OR CI 95%<br>3.461 (1.648–7.269)<br>– Hwang et al. 2013<br>4.044 (1.447–11.301)<br>– Fruscalzo et al. 2014<br>2.236 (0.613–8.161)<br>– Hee et al. 2014<br>Group estimation: 3.346 (1.939–5.775)<br>**Bishop score** | **Successful induction N= 137**<br>**Objective:** to determine the accuracy of cervical elastography in predicting successful induction.<br>**Conclusion:** Although there is a limited number of included studies and heterogeneity in the methods used, cervical elastography appears to be a promising tool for predicting successful labor induction and vaginal delivery in women undergoing medical induction of labor. |
|  |  |  | OR CI 95%<br>0.576 6 (0.282–1.176)<br>– Hwang et al. 2013<br>14.4 (1.754–111.767)<br>– Fruscalzo et al. 2014<br>0.908 8 (0.253–3.251)<br>– Hee et al. 2014<br>Group estimation: 1.455<br>(0.330–6.408) |  |
| Sonoelastography as method for preliminary evaluation of uterine cervix to predict success of induction of labor (25)<br>**2014** | **MULTIVARIABLE LOGISTIC REGRESSION ANALYSIS** | **N=53** | **Logistic regression for predicting cesarean delivery**<br>**EI 4**<br>p=0.0148<br>OR=7.5980<br>(1.4866–38.8330)<br>CI 95%<br>**EI 5**<br>p=0.0321<br>OR=16.628<br>(1.2714–217.481)<br>CI 95%<br>**Nulliparous**<br>p=0.0308<br>OR=0.1171<br>(0.0167–0.8199)<br>CI 95%<br>**Cervical Length**<br>p=0.0461<br>OR=2.6394<br>(1.0168–6.8517)<br>CI 95%<br><u>**ELASTOGRAPHIC INDEX CLASSIFICATION**</u><br>The sonoelastograms were classified into five elastographic index categories based on the prevalence of colors in the cervical tissue:<br>**EI1:** Prevalence of red indicating soft tissue<br>**EI2:** Red-orange<br>**EI3:** Yellow-green<br>**EI4:** Green-blue<br>**EI5:** Prevalence of blue tissue indicating hard tissue | **66.4% spontaneous delivery**<br>**33.96% cesarean section**<br>**N=4 no response to induction**<br>**N=3 fetal distress**<br>**N=6 cervical dystocia**<br>**N=5 mechanical dystocia**<br>The multivariable logistic regression analysis confirmed that **EI4 or EI5, nulliparity**, and higher cervical length values were predictive of cesarean section, while **age** and lower **EI values (1-3)** were not predictive of the occurrence of the event.<br>**EI 1-3 P 82.75% C 17.25%**<br>**EI 4-5 P 45.8% C 54.16%**<br>**Objective:** To determine the sensitivity of sonoelastography in the evaluation of the cervix to predict the success of induction.<br>**Conclusion:** The results of our study indicate that sonoelastography is an innovative technique that could likely allow for a more objective preliminary assessment of the cervix before labor induction. |

By observing the wide range of variables used in all the studies it can be seen how all studies aim to predict the success of labor induction based on the stiffness and cervical length of the cervix. However, to quantify these variables, each study uses its measurements, reflecting a heterogeneous variety regarding the study of cervical stiffness. To study the stiffness of the cervix, shear wave elastography is the device in use. For this purpose, a wide variety of measurement units are employed: Shear Wave Speed (SWS) in meters per second (m/s) [20], Cervical Shear Wave (CSW) measured in kilopascals (Kpa) [18], Cervical Tissue Strain (TS) [17], Angle of Progression (AOP) [21], Electrographic Score [21], Young’s Modulus: used to predict cervical dilation time and the risk of prolonged dilation after labor induction [22], Hardness Ratio (HR) [23], and Elastography Index, which is primarily used to predict the success of labor induction when using oxytocin [19]. The areas where cervical stiffness is measured are the internal and external cervical os. The basis for studying cervical rigidity is to measure the speed at which shear waves propagate through cervical tissue, indicating stiffness, the greater the propagation speed, the stiffer the tissue. A rigid tissue is considered immature, as it is less prepared to dilate at the time of labor.

According to the previous statements, the lower the SWS/SWE, the softer and therefore more mature the tissue, which is more susceptible to indicate a successful labor induction. Therefore, patients with higher SWS/SWE have a greater risk of induction failure, either by not reaching the active phase of labor or by the need for cesarean delivery. In contrast, women with lower SWS/SWE have a higher success rate in labor induction. Tissue strain is greater in women who succeed in induction compared to those who do not achieve a successful induction.

To study the cervical length of the cervix, a transvaginal ultrasound is used. All studies use cm or mm as units of measurement. The anatomical study site is the total length of the cervix, from the internal cervical to the external cervical. Although the length of the cervix varies in each woman, generally, shorter cervices tend to show greater maturity and capacity for dilation than longer cervices at the time of labor induction. Patients with a longer cervix have a higher risk of labor induction failure, either by not reaching the active phase of labor or by the need for cesarean delivery. Conversely, women with shorter cervices have a higher success rate in labor induction. The different measurement units employed in some of the selected articles are reflected in Table 3.

**Table 3:** Summary of the units of measurement used in some of the selected articles.

| ESTUDIO/AÑO | AUTHORS | Stiffness of the uterine cervix |  |  | Cervical length (mm) |  |  |
| --- | --- | --- | --- | --- | --- | --- | --- |
|  |  | Vaginal delivery | Failure | p-value | Vaginal delivery | Failure | p-value |
| Preliminary Results on the Preinduction Cervix Status by Shear Wave Elastography (20)<br>2022 | Jorge Torres, María Muñoz, María Del Carmen Porcel, Sofía Contreras, Francisca Sonia Molina, Guillermo Rus, Olga Ocón-Hernández and Juan Melchor | SWS (m/s) |  | <0.01 | **23.9**<br>**(19.0-29.3)** | 30.5<br>(25.9-32.4) | <0.05 |
|  |  | 2.15<br>(1.97-2.30) | 1.89<br>(1.77-2.05) |  |  |  |  |
| The predictive value of cervical shear wave elastography in the outcome of labor induction (18)<br>2020 | Jing Lu, Yvonne Kwun Yue Cheng, Sin Yee Stella Ho, Daljit Singh Sahota, L. L. Hui, Liona C. Poon and Tak Yeung Leung<sup>1</sup> | SWE (kPa) |  | <0.001 | 24 (16-30) | 29 (22-35) | <0.001 |
|  |  | 5.1<br>(4.2-6.0) | 5.8<br>(4.9-7.0) |  |  |  |  |
| Quantitative Elastography of the Cervix for Predicting Labor Induction Success (17)<br>2015 | A. Fruscalzo, A. P. Londero, C. Fröhlich, M. Meyer-Wittkopf and R. Schmitz | Cervical TS |  | < 0.05 | 30.2<br>(± 8.5) | 32.8<br>(± 3.4) | 0.244 |
|  |  | 0.6 (± 0.1) | 0.8 (± 0.2) |  |  |  |  |
| Successful induction of labor: prediction by preinduction cervical length, angle of progression and cervical elastography (21) 2014 | S. Pereira, A. P. Frick, L. C. Poon, A. Zamprakou and K. H. Nicolades | AOP |  | 0.387 | 19.6<br>(11.7–26.7) | 23.0<br>(15.0–32.2) | 0.049 |
|  |  | 89.1<br>(77.6–98.0) | 88.1<br>(77.4–92.7) |  |  |  |  |
|  |  | Electrographic score |  | 0.187 |  |  |  |
|  |  | 0.266<br>(0.152–0.452) | 0.391<br>(0.184–0.511) |  |  |  |  |
| Quantitative sonoelastography of the uterine cervix prior to induction of labor as a predictor of cervical dilation time (22) 2014 | Lene Hee, Chirtina K. Rasmussen, Jacob M. Schlutter, Puk Sandager & Niels Uldbjerg | Young’s modulus is used, comparing it with the use of the Bishop score and cervical length as superior for predicting cervical dilation time. |  |  |  |  |  |
| Predictive value of cervical length by ultrasound and cervical strain elastography in labor induction at term (23) 2021 | Yimin Zhou, Neng Jin, Qinqing Chen, Min Lv, Ying Jiang, Yuan Chen, Fangfang Xi, Mengmeng Yang, Baihui Zhao, Hefeng Huang and Qiong Luo | HR |  | 0.117 | 24.9<br>(18.1–30.3) | 29.9<br>(24.3–33.8) | 0.005 |
|  |  | 51.42<br>(38.99–62.63) | 59.76<br>(41.47–71.81) |  |  |  |  |
| Elastography of the uterine cervix: implications for success of induction of labor (19) 2011 | M. Swiatkowska-Freund and K. Peris | Elastography index: orificio interno |  | 0.024* | 23.8 ± 7.6 | 26.9 ± 6.4 | 0.12 |

## DISCUSSION

In this systematic review, cervical elastography is shown as an objective, safe, and promising tool to improve accuracy in assessing cervical maturation and, consequently, the success of vaginal induction. Despite the different methods used for cervical elastography, the results remained consistent, plus the conclusions were similar across various studies. Furthermore, the results indicated that cervical elastography is more precise than transvaginal ultrasound measurement of cervical length in predicting successful labor induction and superior to the Bishop score.

Currently, there are two general approaches to imaging tissue elasticity using ultrasound. The first is deformation elastography, which evaluates tissue stiffness through images displayed in a color spectrum, showing how the tissue deforms under pressure. The second approach uses shear waves (SWEI) to assess tissue elasticity; it employs ultrasound beams to create and propagate shear waves through the tissue, with the speed of these waves relating to tissue stiffness. Both approaches provide a quantitative and objective assessment that can replace palpation in clinical practice. In this regard, Torres et al. (2022) [20] evaluates the preliminary capacity of elastography to provide quantitative and objective information about the condition of the cervix based on SWEI, establishing a prognosis for induction failure. These findings are further supported by Fruscalzo et al. (2015) [17], which demonstrated the potential utility of quantitative cervical elastography in predicting induction failure. Additionally, Muscatello et al. (2014) [25], in their multiple regression analysis for predicting cesarean delivery, confirmed that a stiffer cervix, assessed by cervical elastography, along with higher cervical length values, were good predictors for cesarean delivery.

Additionally, this systematic literature review considers multiple variables that influence the prediction of success of a vaginal induction, thereby avoiding cesarean delivery. These variables include parity (multiparous women vs. nulliparous women), BMI, cervical length, Bishop score, cervical elasticity and stiffness, maternal age, and gestational age.

Among all these variables, parity and the Bishop score stand out. The Bishop score is a classic and subjective technique for assessing cervical maturation, routinely used in obstetric clinical practice. Studies such as the one conducted by Lu et al. (2020) [18] affirm through univariate and multivariable analysis for predicting cesarean delivery that the probability of failure in labor induction decreases significantly in multiparous women and increases in women with greater cervical length as evaluated by the Bishop score. In some studies, such as the one by Zhou et al. (2021) [23], it has been observed that the Bishop score may have predictive value for the success of labor induction with a 95% confidence interval. In the systematic review conducted by Hatfield et al. (2007) [15], the Bishop score was compared with cervical length to predict success in induction across 10 studies. There were no significant differences in the diagnostic accuracy of both methods in successful induction, vaginal delivery, active labor, and vaginal delivery within the subsequent 24 hours [14,22]. However, in our review, we consider the subjective nature of the Bishop score, and therefore its difficulty in quantification and low reliability. Consequently, the use of the Bishop score as the sole predictive tool should be approached with caution, highlighting the importance of integrating other objective techniques to improve accuracy in assessing cervical maturation. Although it is a widely used method today due to its simple reproducibility, the Bishop score remains a subjective and imprecise method. Due to its nature, it does not provide us with valuable information and, therefore, should not be the only method for assessing cervical maturation. It is essential to complement it with other objective techniques, such as elastography, to achieve an objective and precise evaluation of cervical stiffness and elasticity.

Currently, there is a growing use of artificial intelligence to improve the effectiveness of assessing cervical maturation in women undergoing induced labor by using data obtained through ultrasound and comparing it with evaluations conducted using the Bishop score. This involves the use of machine learning algorithms such as XGBoost, CatBoost, or RF (Random Forest), offering precise and objective evaluations of the predictive induction failure from the above-described covariates [30, 31].

Among the limitations found in this review is the scarcity of existing literature when posing our research question in search engines, particularly during recent years. Furthermore, the available scientific evidence predominantly consists of observational studies, making it necessary to implement clinical trials and meta-analyses that reinforce the scientific evidence regarding the prediction of success in vaginal induction using novel, quantitative, and objective techniques such as elastography.

Since our results clearly reflect the value of using elastography and its potential superiority over other traditionally used techniques, like the Bishop score or cervical length measurement, we consider it of utmost importance that elastography be implemented in clinical practice, either by combining it with or gradually replacing other traditional techniques already in use, in order to establish an appropriate protocol for the use of elastography in the same context as these other more subjective and less effective traditional techniques.

The measurement of cervical stiffness through elastography and the assessment of cervical length in predicting the success of labor induction are innovative methods that have proven to be superior to the Bishop score, as they are objective and observer-independent, unlike the Bishop score. However, to determine how cervical stiffness is measured using elastography, while all studies agree that softer tissue favors labor induction and stiffer tissue leads to failure in labor induction, they fail to agree on a common measurement (there is a wide variety of measurement units). In contrast, the measurement of cervical length appears to be more consistent across all studies, with results indicating that shorter cervical lengths favor the success of labor induction. All studies illustrate areas under the curve that demonstrate how the incorporation of elastography and cervical length provides greater predictive value than the Bishop score, showing higher area under the curve, sensitivity, and specificity. In conclusion, this is a growing area of study, whose objectivity has already been demonstrated, but there is no standardized method for measuring tissue stiffness.

It is crucial to increase the number of investigations to strengthen the clinical utility of elastography and find a common measurement unit. In addition to continuing to explore new approaches, as well as the potential integration of artificial intelligence—a field that has already begun to be explored with the use of ultrasound but is still in the very early stages of development—there could be opportunities to incorporate data specific to elastography, which is a completely unexplored area at present. The incorporation of artificial intelligence could represent a breakthrough by replacing more subjective and less precise traditional techniques, such as the Bishop score, with new effective, reproducible, and objective methods, thus improving maternal care in labor induction.

## CONCLUSIONS

The analyzed studies indicate that elastography provides valuable information about the elasticity of the cervix and presents itself as a useful tool in predicting the success of labor induction. Despite the variety of measurement units used in these investigations, the consistency in the findings highlights its potential in clinical obstetrics practice. However, these studies also reveal the complexity and diversity of the factors that influence the labor induction process.

## Data Availability

All data produced in the present work are contained in the manuscript

